# Brain-wide neurotransmitter-specific network involvement determines outcome in glioblastoma

**DOI:** 10.64898/2026.03.23.26348837

**Authors:** Philipp J. Koch, Julia Forisch, Robin Kathri, Benedikt Frey, Florian Brembach, Yahya Zghaibeh, Jan Feldheim, Tom Hornberger, Fanny Quandt, Tim Magnus, Götz Thomalla, Matthias Endres, Michael O. Breckwoldt, Varun Venkataramani, Frank Winkler, Michelle Monje, Ulrich Schüller, Malte Mohme, Lasse Dührsen, Katharina Frank, Stefan Bonn, Richard Drexler, Henrik Heiland, Robert Schulz, Franz Ricklefs

## Abstract

**Importance:** Glioblastoma (GBM) cells integrate into neuronal circuits, and preclinical work implicates multiple neurotransmitter (NT) networks as key drivers of invasion and treatment resistance. Whether the integration of GBM within NT-defined large-scale brain networks conveys prognostic information for overall survival (OS) is unknown.

**Objective:** To determine whether NT-specific network involvement of GBM is associated with OS in patients with newly diagnosed Isocitrate dehydrogenase (IDH)-wildtype(wt) GBM.

**Design, Setting, and Participants:** In this observational multicenter cohort study, we analyzed two independent cohorts of adults with histopathologically confirmed IDH-wt GBM. Cohort 1 included 153 patients treated at the University Medical Center Hamburg-Eppendorf, Germany (2012-2024), and cohort 2 comprised 264 patients from the University of Pennsylvania Health System, USA (2006-2018). Preoperative contrast-enhanced MRI was used to derive individual tumor masks, which were spatially mapped onto normative NT-informed structural connectomes spanning 19 receptor and transporter systems.

**Exposures:** Preoperative contrast-enhancing GBM lesions, quantified as patient-specific involvement scores (0-1) within each NT-defined brain network.

**Statistics:** We used partial least-squares regression for variable selection and multivariable Cox proportional-hazards models alongside regularized logistic regression with out-of-sample prediction, adjusted for age, methylguanine methyltransferase (*MGMT)* promoter methylation, and extent of resection, to test associations between NT-specific GBM network involvement and OS.

**Results:** Across 417 patients in two cohorts, greater GBM involvement within cholinergic networks, defined by normative vesicular acetylcholine transporter (VAChT)-weighted as well as dopaminergic D2 receptor involvement, was consistently associated with reduced OS, independent of age, *MGMT* status, and resection extent. Further, cholinergic network involvement showed the strongest contribution to the prediction models. Other NT networks did not show reproducible prognostic effects across cohorts. Tumor-intrinsic hypomethylation of acetylcholine receptor-associated regions correlated with imaging-based cholinergic network involvement and mirrored its prognostic relevance.

**Conclusion and Relevance:** Tumor integration into neurotransmitter-specific brain networks is an independent predictor of poorer survival in GBM. By combining routine clinical MRI with normative NT-informed connectome data, this approach delineates a novel systems-level marker of tumor aggressiveness and supports cholinergic inhibition as a putative therapeutic target in GBM.

## Introduction

Glioblastoma (GBM) is a highly aggressive and fatal primary brain tumor with a poor prognosis despite maximal treatment. Its defining features include diffuse infiltration, rapid proliferation, and nearly universal recurrence, typically occurring within a few centimeters of the resection margin^1^. While classical models have focused on cellular heterogeneity and genomic instability, recent discoveries have reframed GBM as a brain-wide, circuit-integrated disease rather than a purely local infiltrative process^2^. This conceptual shift has highlighted the contribution of neural network involvement to clinical deterioration and reduced overall survival (OS)^3-6^.

Growing evidence points to complex neuron-glioma interactions across multiple scales, ranging from synaptic coupling at the microscale to disruptions in large-scale functional and structural network interactions^3,7-10^. GBM cells have been shown to establish functional synapses with neurons through glutamatergic^11,12^, GABAergic^13^, and cholinergic signaling pathways^8,10,14,15^. Retrograde tracing studies revealed that GBM cells rapidly integrate into anatomically diverse neuronal circuits, with cholinergic neurons in particular promoting tumor proliferation, invasion, and treatment resistance^10,14^. Furthermore, recent work demonstrated circuit specificity, showing that GBM location determines its impact on hierarchical network signaling, and that local glutamatergic and long-range cholinergic inputs act synergistically to accelerate tumor progression^15^. Importantly, pharmacological or genetic perturbation of cholinergic receptors in tumor cells attenuated glioma growth in preclinical models, highlighting a potential therapeutic relevance^3,8,10,14^.

A central challenge in translating these insights to the clinic is not the strength of the preclinical evidence itself, but the lack of scalable approaches to assess neurotransmitter (NT)-related neuron-glioma interactions in human patients. While experimental models have delineated key neurochemical mechanisms underlying tumor-neuron coupling, directly measuring NT-specific network engagement in vivo across large patient cohorts remains difficult. To address this gap, recent neuroimaging studies have related tumor location to systems-level network properties. For example, Salvalaggio *et al*. demonstrated that GBMs involving regions with high white-matter tract density are associated with shorter survival^13^, and Wei *et al*. showed that structural connectome measures capture invasion patterns extending beyond the focal lesion and predict survival^14^. Additional work has identified alterations in both structural and functional brain networks that influence neurological impairment and clinical outcome^15^. However, whether and how brain-wide, NT-informed networks, central to emerging circuit-based models of GBM biology, can be quantified in patients and linked to clinical outcome has remained largely unexplored.

Recent availability of positron emission tomography (PET)-derived normative NT maps has enabled the integration of neurochemical gradients with anatomical network analyses^16^. For example, the integration of NT-informed connectome data into structural network analyses demonstrated its potential for multimodal systems neuroscience approaches for post-stroke recovery^18,30^. In fact, the amount of damage to dopaminergic brain networks, independent of overall network damage, informed about recovery trajectories over time^18^. We therefore sought to determine whether similar principles could be translated from vascular neurology to neuro-oncology.

In the present study, we investigated the association between GBM lesion distribution, NT-informed large-scale brain networks, and overall survival in two independent cohorts comprising 417 patients. Furthermore, we examined whether imaging-derived measures of GBM involvement within specific NT networks correspond to biological alterations, as reflected by epigenetic receptor methylation levels.

## Results

### Cohorts and clinical data

In total, 1005 clinical and radiological datasets of GBM patients were screened for further analysis: 153 were selected for UKE and 264 for UPENN. In UKE, 161 patients were excluded because their *ex domo* acquired imaging data were no longer available at the time of data screening. Twenty-three datasets displaying multifocal lesions and 13 scans with non-enhancing lesions were excluded from further analysis, as were 3 cerebellar and brainstem tumors. For data quality reasons, 13 UKE datasets were excluded due to slice thickness > 3 mm (n = 9), motion artifacts (n = 3), or a tumor protocol without a post-contrast series (n = 1). Three datasets were discarded due to co-registration failure. In UPENN, 114 datasets were excluded for not meeting the current pathological diagnosis of GBM (in accordance with the WHO classification): 67 for multifocal lesions, one for a single brainstem lesion, and 18 for critical mass effect and anticipated registration inaccuracy. Sixty-eight datasets did not meet minimum data quality requirements for further imaging analysis due to slice thickness >3 mm, the absence of post-contrast series, and head motion artifacts. Another 29 datasets with diffuse radiologic findings (i.e., mixed pachymeningeal and leptomeningeal contrast enhancement, dull contrast-enhancing lesions) were excluded due to unsatisfactory lesion segmentation or concomitant intra- or extra-axial lesions of uncertain pathological diagnosis. Eighteen datasets did not meet the quality standard after co-registration **(Figure 1)**. The clinical characteristics for both cohorts are summarized in **Table 1**. The two groups exhibited similar demographic characteristics, with no statistically significant differences in age, sex, or preoperative Karnofsky Performance Status (KPS). In UKE, 58% were men, and the median age of patients was 64 years (range 23-85). Gross total resection (GTR) or near GTR was achieved in 62.1% of cases. In UPENN, 59% were men, with a median age of 63 years (range 33-88). The UPENN metadata did not provide information on the extent of GTR or near GTR. More than 90% resection extent of the contrast-enhancing tumor was achieved in 62.1% of cases. Both groups exhibited mild preoperative functional impairment, with KPS indices of 82 (±15 SD) in UKE and 85 (± 10 SD) in UPENN. In UPENN, *MGMT* promoter methylation status was reported for 121 datasets, of which 62% were unmethylated and 38% were methylated. Statistical comparisons revealed significant differences in OS, with the UKE cohort showing a lower mean OS of 16 (± 16.6) months compared to UPENN, which had a mean OS of 21 (± 25.9) months (**Table 1**).

**Table 1.** Demographic and clinical data. Baseline demographic and clinical variables for the UKE and UPENN cohorts are summarized. Continuous variables (e.g., age, Karnofsky Performance Status, survival times) are reported as mean ± standard deviation (SD) and compared using two-sample *t*-tests. Categorical variables (e.g., sex, *MGMT* status, therapy scheme, resection extent, GBM subtype) are summarized as percentages and compared between cohorts using χ^2^ tests. *N* indicates the number of patients with available data for each variable per cohort (N_UKE_, N_UPENN_). Asterisks (*) denote statistically significant differences between cohorts.

| Variable | UKE | UPENN | P | N<sub>UKE</sub> | N<sub>UPENN</sub> |
| --- | --- | --- | --- | --- | --- |
| Age (y ± SD) | 63.98 ($\pm 11.19$) | 63.34 ($\pm 10.71$) | 0.566 | 153 | 264 |
| KPS (mean ± SD) | 82.48 ($\pm 15.36$) | 85.41 ($\pm 10.16$) | 0.164 | 153 | 37 |
| Sex (%) |  |  | 0.833 | 153 | 264 |
| male | 57.5 | 59.1 |  |  |  |
| female | 42.5 | 40.9 |  |  |  |
| MGMT Status (%) |  |  | 0.071 | 153 | 121 |
| Unmethylated | 50.3 | 62 |  |  |  |
| Methylated | 49.7 | 38 |  |  |  |
| Therapy scheme (%) |  |  |  | 153 | 0 |
| Best supportive care | 5.2 |  |  |  |  |
| Stupp regimen | 76.5 |  |  |  |  |
| RTX mono | 6.5 |  |  |  |  |
| TMZ mono | 5.2 |  |  |  |  |
| CeTeG regimen | 1.3 |  |  |  |  |
| N.A. | 5.2 |  |  |  |  |
| (Near) GTR (%) | 62.1 | 62.1 | 1.0 | 153 | 264 |
| Resection extent (%) |  |  |  | 153 | 264 |
| GTR | 41.8 | N.A. |  |  |  |
| Near GTR (> 90% CE) | 20.3 | 62.1 |  |  |  |
| PR | 15 | 35.7 |  |  |  |
| Biopsy | 22.9 | N.A. |  |  |  |
| N.A. | 0 | 3.4 |  |  |  |
| Overall Survival (m ± SD) | 16 ($\pm 16.6$) | 21.3 ($\pm 26$) | 0.013* | 153 | 264 |
| Deceased (%) | 65 (16 months $\pm 15$) | 97 (18 months $\pm 18$) | | | |
| Censored (%) | 35 (16 months $\pm 19$) | 3 (100 months $\pm 68$) | | | |

**Figure 1.**
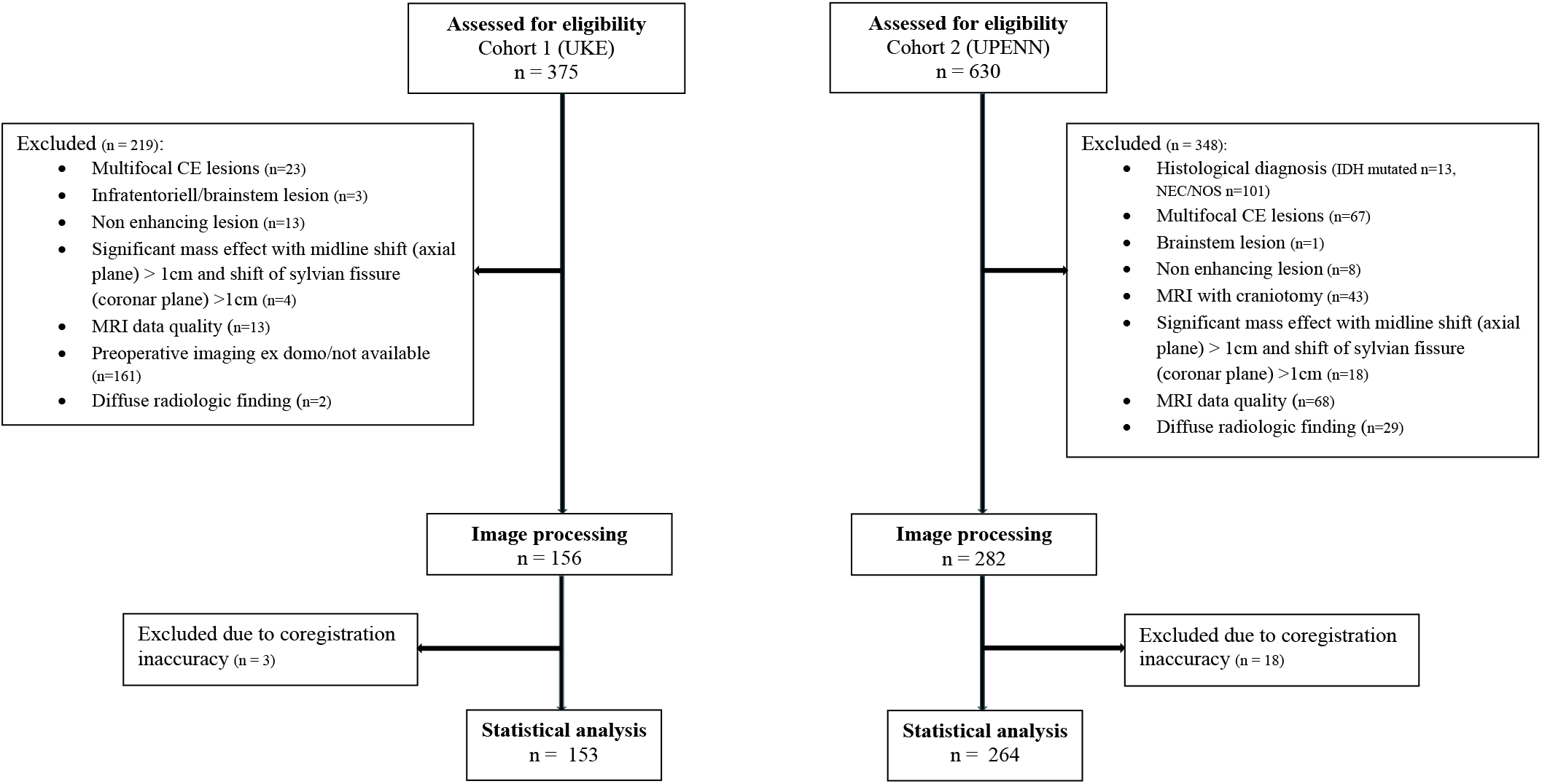
Patient selection for cohort 1 (UKE) and cohort 2 (UPENN). Flowchart illustrating the identification, screening, and inclusion of glioblastoma (GBM) patients across both cohorts. Out of 375 patients initially assessed at UKE, 219 were excluded based on predefined imaging or clinical criteria (e.g., multifocal contrast-enhancing lesions, infratentorial/brainstem involvement, non-enhancing tumors, midline shift > 1 cm, insufficient MRI quality, missing preoperative imaging, or diffuse radiologic findings). After image processing, 3 additional cases were excluded due to co-registration inaccuracies, resulting in 153 patients included in the statistical analysis. Similarly, 630 patients were screened in UPENN database, with 348 excluded due to histological diagnosis inconsistencies, multifocal or brainstem lesions, non-enhancing tumors, craniotomy-related imaging artifacts, midline shift >1 cm, poor MRI quality, or diffuse radiologic findings. Following image processing, 18 cases were excluded due to co-registration inaccuracies, yielding 264 patients for the final statistical analysis.

### Neuroimaging and NT-network involvement

Heatmaps of spatial GBM distribution for both cohorts are shown in **Figure 2A**. Tumors in both datasets were predominantly localized within the temporal and parietal white matter. The spatial distribution of white matter fiber tracts with a strong relation to GBMs shows frequent affection within the posterior limb of the internal capsule, subinsular fibers, parietotemporal white matter, and splenial callosal fibers in both cohorts (**Figure 2B)**. Cortical projections of GBM-related streamlines **(Figure 2C)** converge mainly onto the middle and superior temporal cortex and the temporoparietal junction in both cohorts. **Figure 2D** gives the NT network scores across both cohorts. There was a statistically significant main effect of NT system, with marked differences across systems (F(18, 7488) = 56.99, *P* = 7.7×10^−193^, generalized eta squared = 0.0118), stable after Greenhouse-Geisser and Huynh-Feldt corrections (*P*_GG_ = 4.3 × 10^−33^; *P*_HF_ = 2.5 × 10^−33^). Post-hoc comparisons revealed that 5HTT, 5HT4, 5HT1a, and D1-specific GBM network involvement exceeded the average network involvement, whereas A4B2, MU, NAT, 5HT1b, and NMDA-related network involvement was lower than the average. The GBM relation to DAT, GABA, H3, M1, VAChT, and 5HT2a-related brain networks did not significantly deviate from the grand mean.

**Figure 2.**
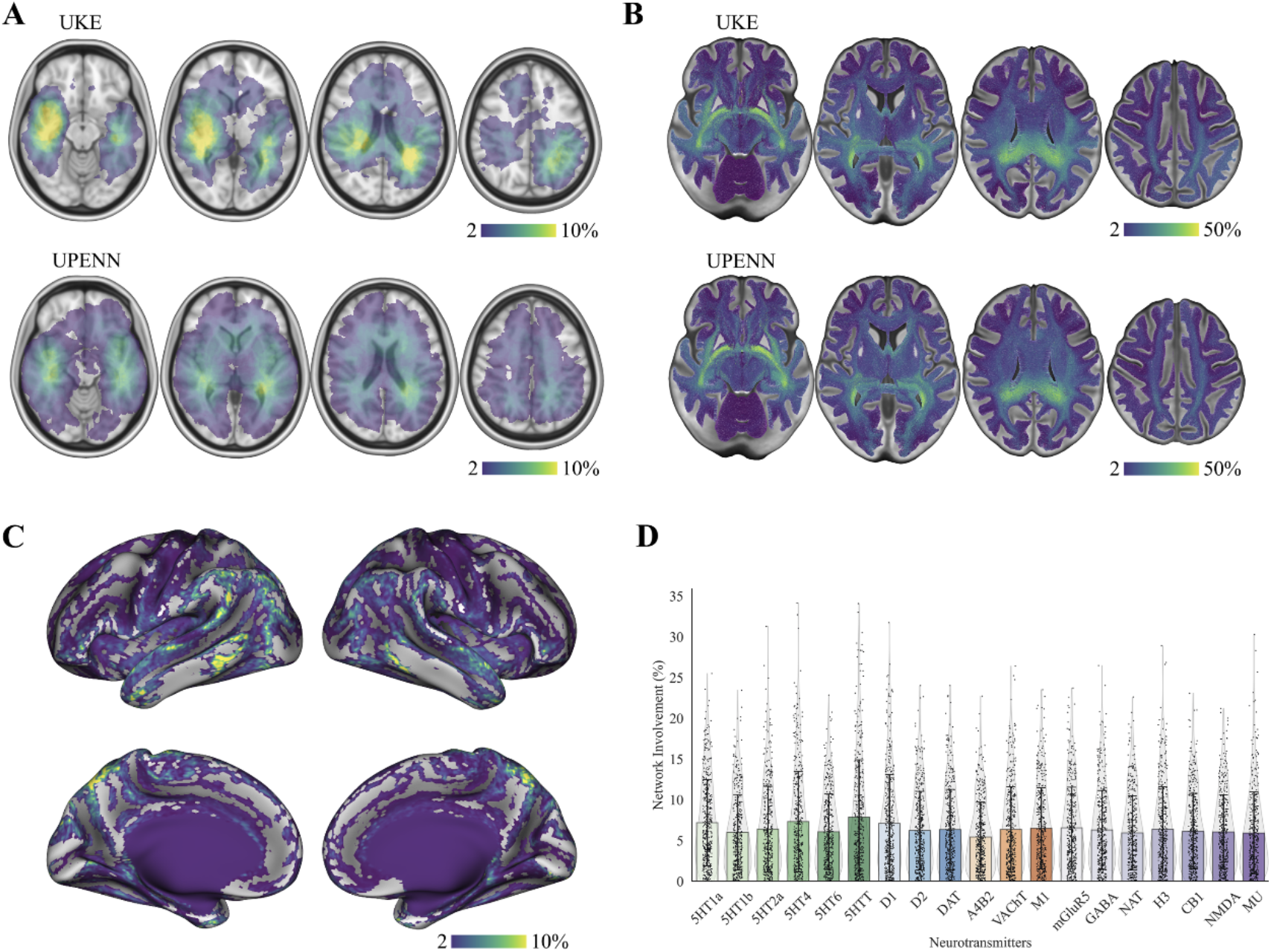
GBM distribution, white matter maps, and regional NT vulnerability across cohorts. **A:** Axial slices in MNI standard brain showing the spatial distribution of GBM localization for the UKE (top) and UPENN (bottom) cohorts. Color scales indicate voxel-wise GBM frequency in terms of the number of patients, demonstrating comparable involvement of periventricular and deep white-matter regions across sites, with a higher frequency in parietotemporal regions. **B:** Group-level structural connectivity maps derived for each cohort. Warmer colors indicate a greater number of streamlines associated with GBM lesions, illustrating convergent patterns of structural network vulnerability across UKE and UPENN. **C:** Cortical surface renderings of streamlines affected by GBM localization across both cohorts. Maps highlight predominant involvement of medial/inferior frontal, temporal, and parietal regions, with consistent patterns across hemispheres. **D:** Bar-and-scatter plots of global scores of NT involvement across both cohorts. Individual dots represent patient-level values; colored bars denote cohort-level means. NT classes are color-coded (serotonergic: green; dopaminergic: blue; cholinergic: orange; other receptor systems: purple).

### Associations between NT-specific network involvement and overall survival

Partial Least Squares (PLS) regression was used to identify the NT-specific network involvement of GBM that was most strongly associated with OS. For UKE, the variable importance in projection (VIP) scores indicated that GBM involvement in D2, DAT, VAChT -informed networks was most strongly associated with OS (**Figure 3A**). For UPENN, GBM involvement in VAChT, NAT, A4B2, 5HT6, D2, CB1-informed brain networks, as well as the overall network affection (Disc-SL), was informative for OS **(Figure 3A)**.

**Figure 3.**
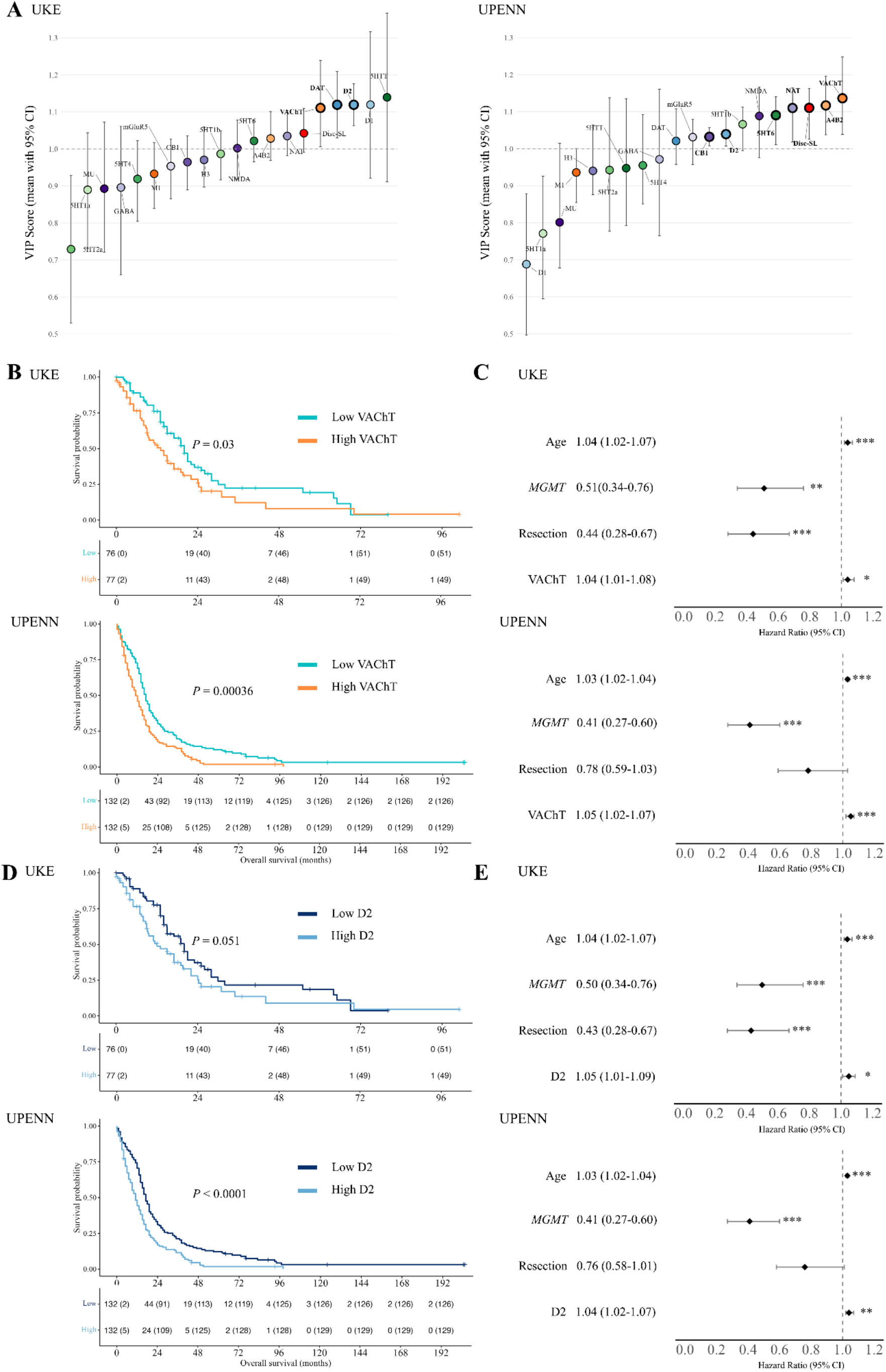
NT-specific network involvement and overall survival in GBM. **A**: Informative predictors for inference of overall survival in UKE (left) and UPENN (right) were selected based on partial least squares (PLS) Cox regression and computation of cumulative variable importance in projection (VIP) scores across three latent components for stability of variable weights and to capture distributed covariance patterns in multivariate data. Subsampling was utilized to estimate 95% confidence interval of VIP scores. Predictors were considered informative when the lower bound of the 95% confidence interval exceeded 1.0 (points with bold circles) and subsequently analyzed using adjusted Cox regression (B-E). For visualization, data were grouped by NT system: green for serotonin, blue for dopamine, orange for acetylcholine, and purple for other NT systems. The global network damage (disconnected streamlines, Disc-SL) is depicted as red. **B:** Kaplan-Meier curves comparing overall survival between patients with high versus low VAChT-specific network involvement (median split) in the UKE (top) and UPENN (bottom) cohorts. In both datasets, greater tumor involvement within VAChT-related networks was associated with shorter overall survival (log-rank *P*-values shown), demonstrating consistent cross-cohort replicability. The tables below the survival curves display, for each time point (in months), the number of patients at risk, with the cumulative number of deaths shown in parentheses, stratified by VAChT expression group (Low vs. High). **C:** Adjusted Cox regression analyses displaying hazard ratios (HRs) and 95% confidence intervals for age, *MGMT* promoter methylation status, extent of resection, and VAChT-specific network involvement in UKE (top) and UPENN (bottom). Established prognostic markers behaved as expected, and VAChT remained an independent predictor of poorer survival after adjustment for covariates in both cohorts. Modelled as a continuous variable ranging from 0 to 100, VAChT involvement exhibited a modest but statistically significant association with reduced overall survival (UKE: HR = 1.04, 95% CI 1.01-1.08, UPENN: HR = 1.05, 95% CI 1.02-1.07). **D:** Kaplan-Meier curves comparing overall survival between patients with high versus low D2-specific network involvement (median split) in the UKE (top) and UPENN (bottom) cohorts. The tables below the survival curves display, for each time point (in months), the number of patients at risk, with the cumulative number of deaths shown in parentheses, stratified by D2 expression group (Low vs. High). **E:** Adjusted Cox regression analyses displaying hazard ratios (HRs) and 95% confidence intervals for age, *MGMT* promoter methylation status, extent of resection, and D2-specific network involvement in UKE (top) and UPENN (bottom). Modelled as a continuous variable ranging from 0 to 100, D2 involvement exhibited a modest but statistically significant association with reduced overall survival (UKE: HR = 1.05, 95% CI 1.01-1.09, UPENN: HR = 1.04, 95% CI 1.02-1.07).

To further interpret the most informative PLS-derived predictors, adjusted multivariable Cox regression models were fitted to examine the association between GBM network involvement within the significant NT systems and OS. All top predictors demonstrated statistically significant associations with OS in both cohorts. At the model level, inclusion of NT-specific network involvement consistently improved model fit relative to baseline covariate models. Across both cohorts, only GBM involvement within VAChT-related networks and D2-related networks showed a robust association with OS, and its inclusion statistically significantly improved baseline model performance **(Supplementary Table 1)**. Specifically, a higher degree of tumor involvement within VAChT-related networks was associated with shorter OS in both datasets, with a 5% increase in mortality risk per 1% network involvement (HR = 1.05, 95% CI 1.02-1.08). Comparable associations were observed for D2-network involvement **(Supplementary Table 1)**.

Kaplan-Meier curves and log-rank tests confirmed a significantly divergent survival trajectory, based on a median split of VAChT and D2 scores, in both cohorts, with patients exhibiting greater or smaller network involvement by GBM **(Figure 3B,D**).

Using the empirically observed median overall survival of 18 months in the lowest VAChT involvement quantile as the reference group, the maximal VAChT network involvement observed in the analyzed cohort (26%) corresponds, under the proportional hazards assumption, to an approximately 3.6-fold increase in hazard, translating into a model-based estimate of median overall survival of roughly 5 months. This represents an absolute reduction in median survival of about 13 months compared with the low-risk group. Accordingly, a 10% increase in VAChT network involvement corresponds to an approximately 1.6-fold increase in hazard and is associated with a model-based reduction in median overall survival from 18 months to approximately 11-12 months, representing an absolute decrease of about 6-7 months. In the Cox regression models, higher age and unmethylated *MGMT* status were consistently associated with worse OS; lower resection extent was additionally associated with poorer OS in the UKE cohort **(Figure 3C)**. Sensitivity analyses were performed exclusively in the UKE cohort owing to data availability. The KPS index did not show a statistically significant association with OS (*P* = 0.11; HR = 0.99; 95% CI 0.97-1.00). When controlling for KPS, the VAChT network involvement did not show a statistically significant association with OS (*P* = 0.122, HR: 1.03, 0.99-1.07). No further radiochemotherapy was associated with worse OS (*P* = 0.0022, HR: 2.23, 1.34-3.74). Correcting for the influence of radiochemotherapy did not challenge the significant effect of VAChT network involvement on OS.

### Out-of-sample prediction

To assess whether NT-informed network damage measures improve individual-level outcome prediction beyond established clinical variables, we evaluated out-of-sample performance in a combined UKE/UPENN dataset. The full model comprising clinical baseline variables together with NT-informed network damage measures and global structural disconnection (Disc-SL) showed consistently better discrimination than the base clinical model alone. As illustrated by the mean ROC curves **(Figure 4A)**, the full model achieved higher true positive rates across most false positive rate thresholds, indicating a modest but systematic improvement in out-of-sample discrimination (AUC mean ± SD: base 0.67 ± 0.05, full: 0.71 ± 0.05). Evaluating the individual predictors’ contribution within the predictive framework, cholinergic network damage measures, in particular VAChT and A4B2, exhibited the largest mean absolute standardized coefficients, indicating strong contributions to the linear predictor when selected, next to known clinical covariates such as age, resection, and *MGMT* status **(Figure 4B)**. In contrast, most other NT-informed measures showed substantially smaller coefficient magnitudes. Selection frequency analysis confirmed that VAChT and A4B2 were among the most consistently retained predictors across resampling iterations, alongside age, resection, *MGMT*, Disc-SL, NAT, and 5HTT involvement **(Figure 4C)**. This underscores the robust prognostic relevance of known clinical covariates and the identified influence of cholinergic network involvement.

**Figure 4.**
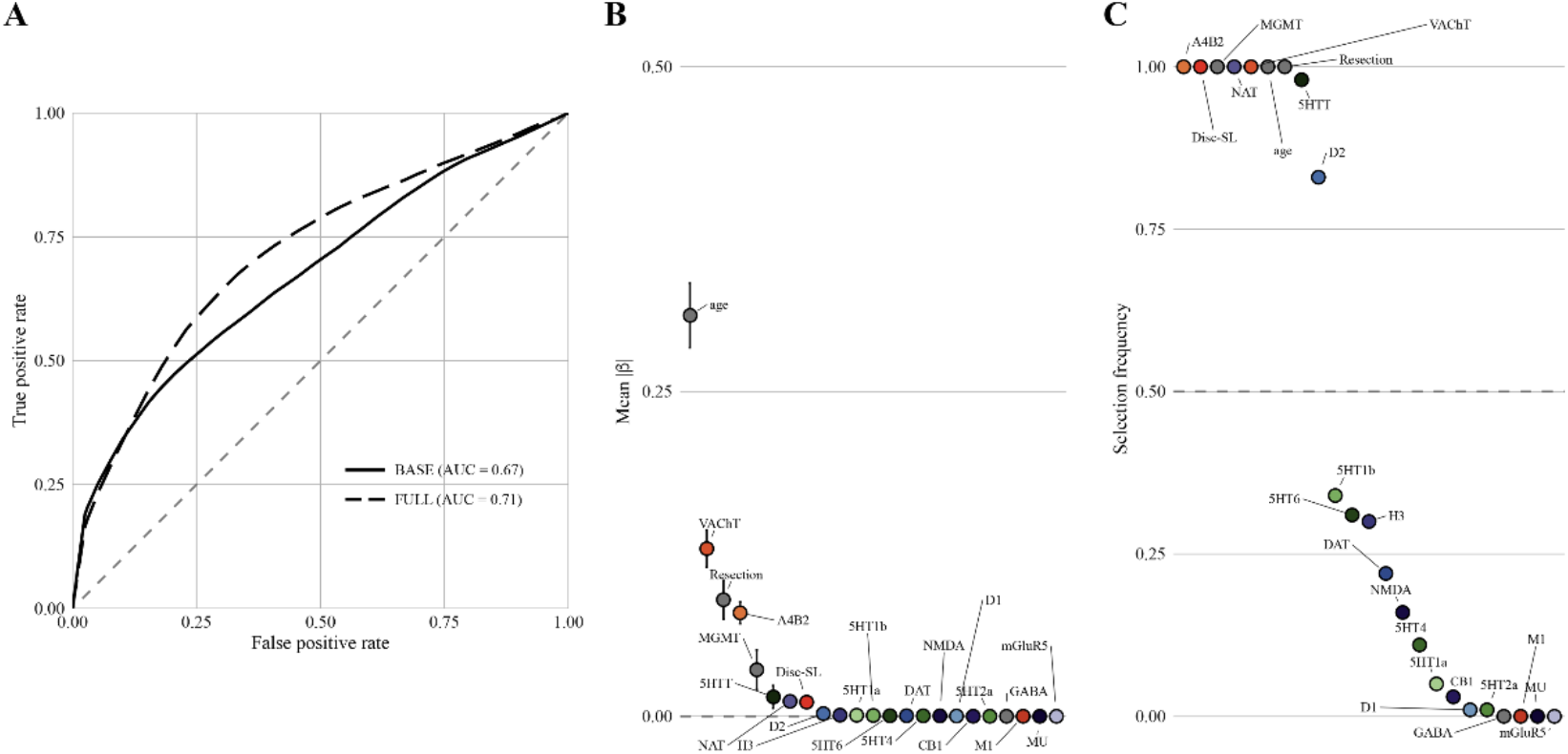
Predictive performance and feature importance of elastic-net models for overall survival. All predictive analyses shown were performed in the combined UKE and UPENN cohorts using an out-of-sample elastic-net modeling framework to evaluate the generalizability of clinical variables and NT-informed network damage measures for dichotomized overall survival (median split). **A:** Receiver operating characteristic (ROC) curves comparing the BASE model (clinical variables only) and the FULL model (clinical variables plus NT-informed imaging features). The dashed diagonal indicates chance-level performance. Area under the curve (AUC) values are reported for each model. **B:** Mean standardized regression coefficients (β) of the FULL elastic-net model, averaged across repeated stratified cross-validation folds. Points represent standardized mean β estimates, and vertical lines indicate variability across repetitions. Positive values indicate an association with increased hazard. **C:** Feature selection frequency across repeated elastic-net models. Each point denotes the proportion of cross-validation repetitions in which a feature was selected (non-zero coefficient). The dashed horizontal line marks a selection frequency of 0.5. Clinical variables and NT-related features are color-coded by system.

When evaluating adjusted Cox regression models restricted to patients whose tumors overlapped predefined anatomical regions of interest, significant model improvement in the association between VAChT-specific network involvement and OS was observed only in GBMs located within the prefrontal cortex (PFC), particularly the ventromedial PFC (vmPFC) **(Supplementary Table 2)**.

### Overall survival by MGMT status and VAChT involvement

Given the robust effect for VAChT involvement in correlative and predictive modelling, we next explored the combined effects of VAChT and MGMT promotor methylation on OS in a joint analysis of the UKE and UPENN cohorts, Kaplan-Meier curves demonstrated statistically significant differences in OS between *MGMT* promoter, methylated and unmethylated tumors **(Figure 5A)**, as well as between groups defined by a median split of VAChT involvement **(Figure 5B)**. When patients were stratified jointly by *MGMT* status and VAChT network involvement, survival curves diverged markedly, with the poorest outcomes observed in patients with an unmethylated *MGMT* promoter and high VAChT network involvement **(Figure 5C)**.

**Figure 5.**
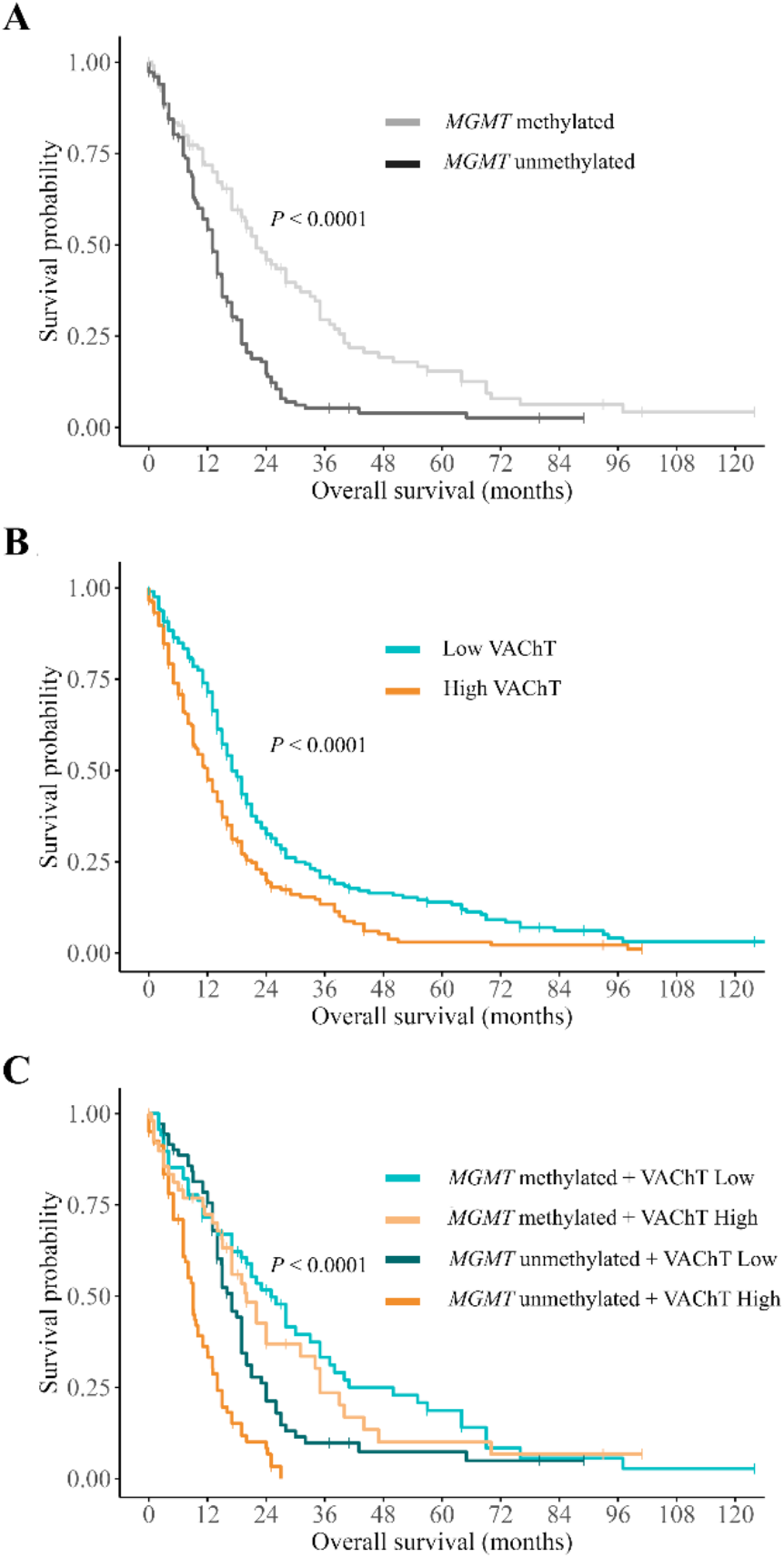
Kaplan-Meier survival analyses stratified by *MGMT* status and VAChT network involvement. All survival analyses shown were performed in the combined UKE and UPENN cohorts, assessing the effects of *MGMT* promoter methylation, VAChT network involvement, and their combined influence on overall survival. *P*-values were derived from log-rank tests. **A:** Overall survival according to *MGMT* promoter methylation status, demonstrating significantly prolonged survival in patients with *MGMT*-methylated tumors compared with *MGMT*-unmethylated tumors. **B:** Overall survival stratified by VAChT network involvement (median split), showing shorter survival in patients with high VAChT involvement compared with low VAChT involvement. **C:** Combined stratification by *MGMT* status and VAChT network involvement, illustrating a graded separation of survival curves with the most favorable outcome in *MGMT*-methylated tumors with low VAChT involvement and the poorest outcome in *MGMT*-unmethylated tumors with high VAChT involvement. Overall survival is shown in months.

### Genome-wide DNA methylation

To examine whether the survival association observed for imaging-based cholinergic network involvement is reflected at the level of tumor-intrinsic epigenetic regulation, we analyzed genome-wide DNA methylation data from a cohort of 362 glioblastoma tumors^3^, focusing on genes related to NT signaling. Across most NT systems, CpG sites annotated to NT-related genes showed minimal interindividual variability and were predominantly unmethylated, precluding meaningful stratification into methylated and unmethylated groups. Consequently, integrated imaging- and survival-based analyses were not feasible for most NT-associated genes. To specifically assess postsynaptic components of cholinergic neuron-glioma interactions, we next examined the methylation status of cholinergic receptor genes. CpG sites annotated to muscarinic acetylcholine receptor genes exhibited near-complete demethylation across tumors **(Supplementary Figure 1)**, preventing informative patient stratification. In contrast, CpG regions annotated to nicotinic acetylcholine receptor genes displayed sufficient methylation variability to permit further analysis. Notably, methylation levels within these nicotinic receptor-associated regions correlated with imaging-based measures of cholinergic network involvement. Hypomethylation of nicotinic receptor-associated CpG sites was strongly correlated with VAChT-based network involvement (Spearman ρ = −0.29, *P* = 0.0004, n = 149) (**Figure 6A**). Consistently, tumors classified as having high imaging-based VAChT network involvement exhibited significantly lower methylation levels in nicotinic receptor-associated regions than tumors with low VAChT involvement (**Figure 6B**). Stratification of tumors by methylation status of nicotinic cholinergic receptor-associated regions revealed a significant difference in overall survival (*P* = 0.0075), supporting the concordance between imaging-derived cholinergic network involvement and tumor-intrinsic epigenetic regulation as biologically relevant contributors to patient outcome (**Figure 6C**). Together, these findings support a conceptual dissociation between presynaptic and postsynaptic components of neuron-glioma communication: while NT-informed connectome maps primarily capture presynaptic cholinergic network integration at the brain-wide level, epigenetic variation within the tumor appears biologically informative only for specific postsynaptic receptor classes, most notably nicotinic acetylcholine receptors.

**Figure 6.**
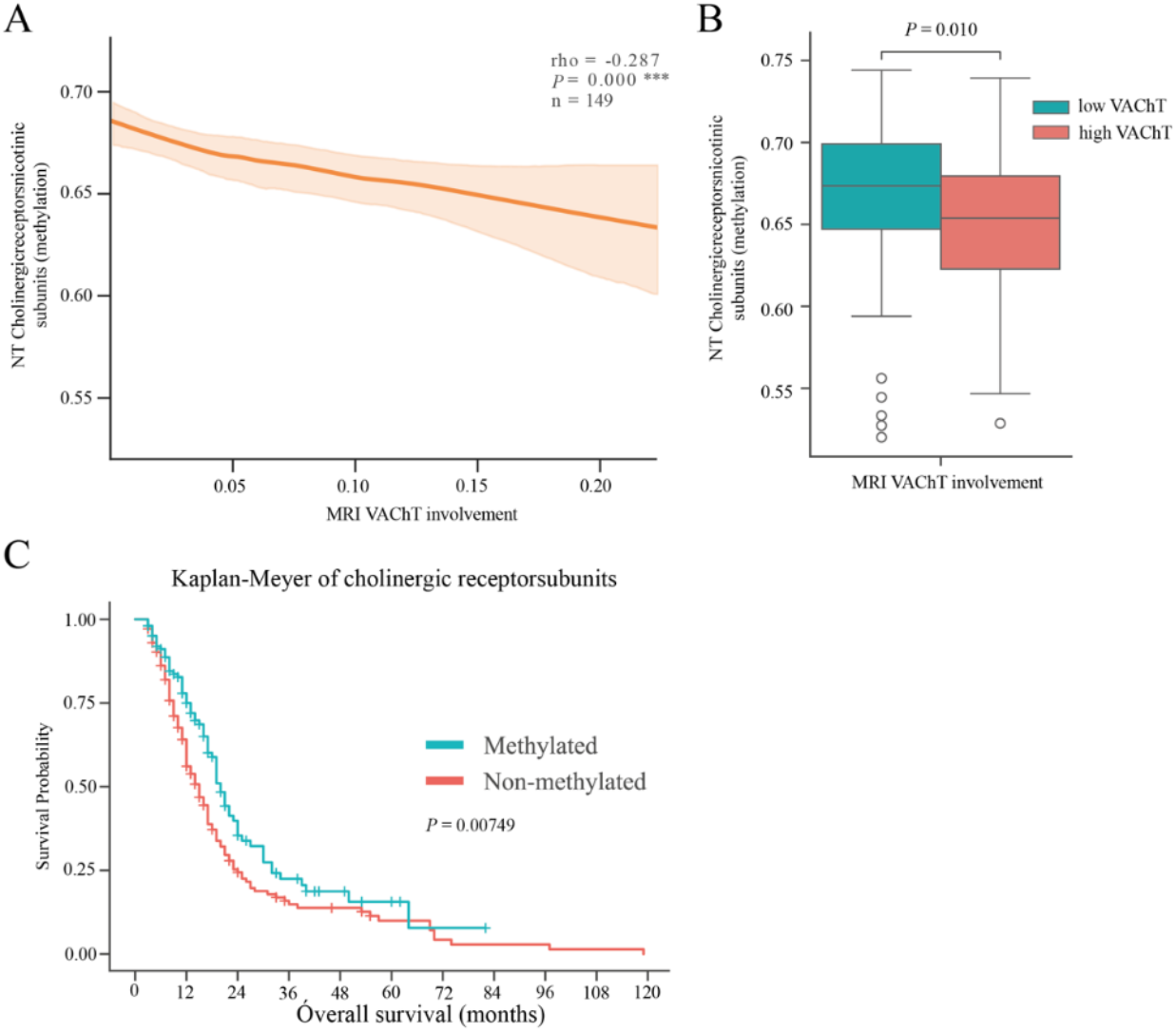
Imaging-based cholinergic network involvement is reflected at the level of tumor-intrinsic epigenetic regulation. DNA methylation profiles were done on the UKE cohort and 149 patients were analysed. **A:** Greater VAChT network involvement is significantly associated with hypomethylation of cholinergic nicotinic receptor subunits (Spearman’s ρ = −0.29, P = 0.0004, n = 149). **B:** Median stratification of VAChT network involvement demonstrates a significant difference in methylation status of cholinergic nicotinic receptor subunits in GBM tumors. **C:** Kaplan-Meier analysis reveals a significant difference in overall survival between methylated and unmethylated tumors, with unmethylated tumors associated with reduced survival

## Discussion

The bidirectional involvement of glioma cells in neuronal networks has recently been identified as a major factor in tumor progression^3,7-10,12^. Augmenting MRI with NT-network information improves prediction of tumor growth and clinical outcome, supporting a pathophysiological role of NT systems in GBM progression. Here, we show that NT-informed structural brain networks provide a principled framework for quantifying long-range network engraftment in GBM and present the following findings: 1) Across two large, independent cohorts, GBMs showed consistent spatial overlap with temporal-parietal white matter and convergent infiltration of major cortico-subcortical pathways. 2) NT-specific network involvement varied substantially in GBM. 3) PLS analysis, Cox regression, and out-of-sample prediction via elastic nets identified multiple NT-specific networks whose involvement in GBM was associated with overall survival (OS) in both cohorts, with the brain-wide cholinergic network biased towards the expression of VAChT emerging as the most robust and reproducible predictor. 4) Higher VAChT-specific network involvement was independently associated with shorter OS, statistically significantly improving model performance, producing clearly divergent Kaplan-Meier survival trajectories, and improving predictive modeling for OS. 5) The prognostic influence of VAChT-specific network involvement remained statistically significant across multivariable and region-restricted analyses, particularly for tumors affecting the prefrontal and ventromedial prefrontal cortex. 6) Concordant associations of VAChT-weighted cholinergic network involvement and nicotinic acetylcholine receptor hypomethylation provide systems-level and tumor-intrinsic evidence for biologically relevant cholinergic neuron-glioma interactions. These findings are in line with previous preclinical studies suggesting that NT-network involvement mediates tumor growth and invasiveness^7-10,12^. We provide clinical indications, supported by prior preclinical studies, that acetylcholine plays a pivotal role in neuron-glioma synaptic communication^8,10^. This underscores the potential of neurochemical pathways as novel therapeutic targets in GBM. Moreover, our results highlight the possibility of developing imaging-based biomarkers to non-invasively estimate tumor aggressiveness and prognostic outlook based on the tumor’s chemical network integration.

Using transsynaptic tracing, various groups have demonstrated that human GBM cells rapidly integrate into both local and long-range neuronal circuits through complex interactions with multiple environmental neuromodulatory systems^14^. Single-cell sequencing analyses revealed high expression of a broad spectrum of NT receptor classes in GBM^10^, suggesting that tumor cells are primed to engage with neuromodulatory networks. Among these, direct glutamatergic and cholinergic inputs most prominently foster tumor progression into the neuronal connectome of brain tumors. Yang *et al*. systematically investigated such inputs and found that cholinergic projections onto GBM cells were the most consistently observed, predominantly originating from the basal forebrain, including the diagonal band of Broca^15^. Accumulating evidence indicates that acetylcholine enhances GBM cell motility and migration via metabotropic CHRM3 receptors, thereby accelerating tumor invasion and reducing survival^17^. Tetzlaff *et al*. identified putative cholinergic synapses in human GBM tissue and patient-derived xenograft models and demonstrated that acetylcholine, ATP, glutamate, and dopamine elicit strong calcium responses in GBM cells^10^. Pharmacological blockade of muscarinic acetylcholine receptors markedly reduced acetylcholine-induced calcium events, and high CHRM3 expression correlated with higher invasiveness and neural scores^3,10^. Knockdown of CHRM3 significantly attenuated cortical tumor expansion in xenograft models, collectively identifying acetylcholine as a key mediator of neuron-tumor connectivity and GBM progression. Beyond cholinergic signaling, preclinical studies show that inhibition of neuronal activity, particularly AMPA-mediated glutamatergic transmission, combined with radiotherapy, enhances therapeutic efficacy, highlighting glutamatergic neuron-glioma communication as another major driver of GBM invasiveness^10^.

Because synaptic connections can facilitate GBM migration along axonal pathways, NT-specific white-matter networks provide a biologically grounded framework for assessing GBM progression within brain connectomes, even in clinical cohorts. Our approach leverages routine clinical imaging to enable in vivo characterization of neuron-tumor interactions by mapping GBM infiltration within structural connectomes enriched for distinct NT-profiles^18^. Using population-based structural connectivity weighted by regional NT-specific expression patterns, we quantify the extent to which GBM involves, or, at a speculative note, might integrate into neurochemically defined large-scale brain networks (**Figure 2**). We demonstrate that greater tumor overlap within cholinergic networks, defined by structural connectivity to regions with high vesicular acetylcholine transporter (VAChT) expression, is associated with shorter overall survival. These findings extend preclinical evidence that the tumor microenvironment’s neuromodulatory landscape shapes GBM progression along specific long-range neurochemical pathways and provide in vivo confirmation of this mechanism across two independent clinical cohorts. Patients whose tumors show stronger engagement within VAChT-defined networks may harbor more invasive tumors, reflecting enhanced integration into cholinergic circuits known to promote motility and infiltration, worsening prognosis. Notably, the strongest association between VAChT-specific network involvement and survival was observed in tumors located within the prefrontal cortex, aligning with experimental data indicating that cholinergic inputs arise predominantly from basal forebrain projection systems^15^.

As an additional finding in the correlative statistical approach, we also observed that, across both cohorts, greater integration of tumor extent into D2-defined networks was associated with shorter OS. In fact, experimental studies indicate that D2 receptor signaling promotes glioblastoma survival, proliferation, and invasion^31-34^. Consistently, clinical data link higher D2 expression to poorer survival, and D2 antagonists suppress glioblastoma growth in vitro and in vivo^35,36^. Importantly, the dopamine D2 receptor antagonist ONC201 (dordaviprone) has demonstrated antitumor activity in glioma models and early clinical studies^37-39^ and is currently being evaluated in clinical trials for gliomas, including the phase III ACTION trial in H3K27M-mutant diffuse midline glioma^40^. Although our findings support an association between D2-network involvement and survival, the present predictive analyses did not indicate a pronounced effect, suggesting that the observed association may not reflect a strong clinical impact or heterogeneous effects in need of further classification.

Building on the robust imaging-based association between VAChT-defined cholinergic network involvement and OS across both correlational and predictive analyses, our methylation findings provide a complementary tumor-intrinsic layer that is consistent with a presynaptic-postsynaptic framework of neuron-glioma communication. Specifically, hypomethylation within CpG regions annotated to nicotinic acetylcholine receptor (nAChR) genes was associated with higher imaging-derived cholinergic network involvement (most prominently VAChT) and was tracked with shorter survival. Mechanistically, these findings align with converging evidence that glioblastoma cells integrate into local and long-range neural circuits, including basal forebrain cholinergic inputs, and that such neuron-to-tumor coupling can promote proliferation, invasion, and treatment resistance^10,13-15^. Recent work has emphasized that neuromodulatory afferents, beyond glutamatergic synapses, form part of a brain-wide neuron-glioma connectome, with cholinergic activity and acetylcholine signaling emerging as functionally relevant drivers of glioma progression in multiple models^8,10,14^. In this context, nAChR hypomethylation may represent an epigenetic state that increases “postsynaptic” tumor receptivity to cholinergic input, conceptually paralleling the broader paradigm in cancer neuroscience whereby neuronal activity fuels glioma growth through activity-regulated pathways and bona fide synaptic signaling^7,19^. Together, the concordance between presynaptic network-level cholinergic engagement (imaging) and postsynaptic tumor-level epigenetic variation (nAChR-associated methylation) supports the notion that cholinergic neuron-glioma interactions are not only detectable at a systems level but may also be reflected in tumor-intrinsic regulatory programs that are relevant to patient outcome. ^4^.

The present findings suggest several potential clinical applications. First, quantifying GBM involvement within VAChT-defined brain networks may serve as a novel prognostic imaging biomarker, complementing established markers such as age, *MGMT* status, or the Karnofsky Performance Score, in estimating individual patient outcomes. The prognostic impact of VAChT network involvement was comparable to that of age at the unit level, one of the most important established predictors of outcome in glioblastoma, with hazard ratios of 1.05 per 1% VAChT involvement and 1.03 per year of age, given that both variables operate over comparable numeric ranges. Increasing VAChT involvement translated into clinically meaningful risk differences, with model-based estimates indicating that a 10% increase was associated with a reduction in median overall survival from 18 months to approximately 11-12 months. While *MGMT* methylation showed a strong protective association with OS (HR = 0.41-0.51), an increase of approximately 14-18% in VAChT involvement conferred a comparable change in hazard, and across the full observed range of VAChT involvement (0-26%), the associated hazard increase exceeded that of *MGMT*. In predictive modeling, the contribution of VAChT network involvement, as reflected by standardized beta coefficients, was intermediate between that of age and MGMT or resection level. Considering the combined effects of both *MGMT* and VAChT characteristics, the most favorable outcome was observed in *MGMT*-methylated tumors with low VAChT involvement, whereas patients with an unmethylated *MGMT* promoter and high involvement within VAChT-related networks exhibited the poorest survival. This finding underscores the clinical relevance of cholinergic network integration as an independent, biologically meaningful, additive/synergistic prognostic factor for mortality in glioblastoma. Validation in larger, multicenter cohorts and through predictive modeling will be essential to confirm the prognostic utility of NT-specific network measures in GBM. Second, our results strengthen preclinical evidence implicating cholinergic modulation in glioma invasiveness, thereby opening avenues for targeted neuromodulatory therapies. Pharmacological strategies, such as anticholinergic agents (e.g., Biperiden or Trihexyphenidyl^41^), or neuron-targeted approaches, including viral vectors that silence or ablate connected neurons, may influence tumor progression, with therapeutic efficacy potentially depending on the degree of glioma integration within cholinergic circuits. Emerging evidence indicates that the therapeutic benefit of D2 receptor antagonists in glioblastoma may be heterogeneous across patients. The presented algorithm could support patient stratification based on NT-specific network involvement and help estimate the likelihood of response to such interventions. Ultimately, randomized controlled trials will be necessary to evaluate the clinical impact and therapeutic potential of anticholinergic or neuromodulation-based strategies in GBM.

Although non-enhancing perifocal FLAIR alterations are thought to reflect diffuse tumor infiltration along white-matter pathways and may critically contribute to distant recurrence and the overall incurability of GBM, the imaging quality of our raw data did not permit reliable analysis of this lower-grade infiltrative component. Patients with large tumors causing substantial midline shift were not included, which may introduce selection bias and limit the generalizability of the findings to less space-occupying lesions. Interestingly, our study did not replicate prior reports suggesting a strong glutamatergic influence on GBM progression. Earlier studies have linked excessive glutamate release to GBM migration along neuronal pathways^10^, a process that may not be adequately captured by our contrast-enhancing tumor masks. This limitation likely reflects our focus on the contrast-enhancing tumor core, which represents the high-grade compartment characterized by gadolinium accumulation due to blood-brain barrier disruption^42^. Additionally, the NT-specific network maps used to assess glutamatergic involvement in our analysis were derived from distributions of NMDA and mGluR5 receptors. In contrast, preclinical work has predominantly implicated AMPA receptors as the key mediators of neuron-glioma synaptic interactions. This mismatch may explain the lower specificity observed in our dataset for glutamatergic networks. Future studies should incorporate AMPAR-specific connectivity maps to more accurately characterize the contribution of glutamatergic neuromodulation’s contribution to GBM progression in clinical cohorts.

Finally, NT-informed networks were derived from normative, population-based atlases^16^ rather than patient-specific molecular imaging. Consequently, our approach does not account for potential circuit remodeling or neurotransmitter reorganization induced by GBM itself, as suggested by recent evidence of large-scale neural circuit rewiring in GBM^4^. Importantly, however, the use of normative NT maps represents a widely adopted strategy in human systems neuroscience^43^, including stroke recovery^30,44^, healthy aging^45^, and Parkinson’s disease and epilepsy^16,46^. Thus, although normative atlases cannot capture tumor-induced, patient-specific remodeling of NT systems, they provide a biologically grounded systems-level scaffold that has proven informative across multiple neurological conditions. Future studies combining individual PET-based NT imaging with tumor-specific network mapping will be required to refine and validate the present findings at a personalized circuit level.

## Conclusions

In this study, we demonstrate, across two independent clinical cohorts, that GBMs extending into large-scale neurotransmitter specific, particularly cholinergic, brain networks are associated with statistically significant poorer overall survival. These findings are in line with preclinical evidence suggesting that cholinergic modulation promotes tumor motility and progression. Clinically, our results support the exploration of novel neuromodulatory treatment strategies in GBM. In addition, quantifying tumor involvement within NT-specific networks may provide a valuable prognostic biomarker for overall survival and may ultimately help guide individualized therapeutic interventions.

## Participants & Methods

### Cohorts and clinical data

Two independent cohorts were analyzed. For cohort 1, radiographic and clinical data from 375 patients undergoing surgery for isocitrate dehydrogenase (IDH)-wild-type GBM were identified from a prospectively collected and maintained database of the Department of Neurosurgery at the University Medical Center Hamburg-Eppendorf, Germany (abbreviated as UKE). Informed written consent was obtained from all patients and experiments were approved by the medical ethics committee of the Hamburg chamber of physicians (PV4904). Patients were included if they received adjuvant combined radiochemotherapy (following Stupp^20^ or CeTeG^21^ regimens), monotherapy with temozolomide, radiotherapy, or best supportive care. Informed written consent was obtained from all patients. Diagnosis was based on the current WHO classification of central nervous system tumors (WHO 2021^22^). The extent of resection was stratified into gross total resection (GTR), partial resection (PR), and stereotactic biopsy^3,23,24^. A GTR was defined as a complete removal of contrast-enhancing parts, a near GTR as a removal of more than 90 % of the contrast-enhancing parts (including^24^), whereas a resection of less than 90 % was defined as PR (including RANO^24^). The extent of resection of contrast-enhancing regions was evaluated on MRI up to 48 hours after surgery. Overall survival (OS) and progression-free survival (PFS) were measured from the date of surgery until death from any cause or progression^25,24^. For cohort 2, data from the database of the University of Pennsylvania were used (https://www.cancerimagingarchive.net/collection/upenn-gbm, abbreviated as UPENN). Six hundred thirty imaging datasets, along with available clinical metadata, were screened for further processing. In both cohorts, only patients with IDH wild-type GBM and a unifocal supratentorial lesion were included. Datasets with craniotomies were excluded to rule out recurrent tumor lesions. To achieve satisfactory co-registration results, lesions with significant mass effect were excluded (midline shift on axial plane >1cm, as well as shift of the Sylvian Fissure (coronar plane >1cm). To obtain accurate lesion segmentation, imaging data with slice thicknesses greater than 3mm were discarded.

### Imaging process and NT-informed network assessment

To quantify the overlap of GBM and NT-informed structural brain networks, JF and PJK manually delineated binary GBM masks from T1-weighted images. Contrast agent enhancement was chosen as the spatial extent of the GBM. Binary masks were transformed into Montreal Neurological Institute (MNI) standard space. The quality of frontal sulcus, sylvian fissure, lateral brain border). If fewer than eight landmarks were correctly aligned, this patient was excluded from further analyses due to misregistration. Transformed masks were then integrated into normative NT-informed structural connectome data. In brief, 19 different normative NT-receptor and transporter density maps, i.e., positron-emission tomography (PET) tracer maps for serotonergic, dopaminergic, cholinergic, glutamatergic, GABAergic, and noradrenergic systems, were obtained from Hansen et al.^16^. Each tracer map was co-registered to an averaged Human Connectome Project-derived normative connectome with 2 million streamlines (SL)^16^. Each streamline was assigned the product of specific NT densities in its endpoints, defined as the NT-specific streamline weight (SL_NTW_). To allow for comparability between different NT systems, SL_NTW_ data were normalized to the sum of weights across all SL within each NT map. Thus, 19 new normative NT-informed connectivity maps in MNI space were generated, publicly available on GitHub (https://github.com/phjkoch/NTDisconn). Higher SL_NTW_ values within these connectomes indicate a specific SL that connects brain areas with higher NT receptor and transporter densities, respectively. After integrating binary GBM masks into these connectivity maps, we selected all streamlines associated with the GBM masks and summed their respective SL_NTW_ scores. After normalization, the estimate of NT-specific network involvement ranges from 0% to 100% involvement. Aside from the NT-specific network involvement, we also computed the unbiased sum of SL as a measure of the overall network damage due to the GBM lesion (abbreviated as Disc-SL). To assess the anatomical topography of GBM, tumor masks were also overlaid with anatomical regions of interest, including the hippocampus, lentiform nucleus, ventromedial prefrontal cortex (vmPFC), and prefrontal cortex (PFC), anatomical regions associated with high glioma incidence^47,48^. Anatomical landmarks were derived from the Brainnetome Atlas^49^.

### *Methylation* analyses

DNA was extracted from tumors and analyzed for genome-wide DNA methylation patterns using the Illumina EPIC (850k) array. Processing of DNA methylation data was performed with custom approaches^50^. Methylation profiling results from the first surgery were submitted to the molecular neuropathology (MNP) methylation classifier v12.8 hosted by the German Cancer Research Center (DKFZ)^50,51^. Patients were included if the calibrated score for the specific methylation class was >0.84 at the time of diagnosis in accordance with recommendations by Capper et al.^50^. For IDH-wildtype GBM, patients with a score below 0.84 but above 0.7 with a combined gain of chromosome 7 and loss of chromosome 10 or amplification of epidermal growth factor receptor (*EGFR*) were included in accordance with cIMPACT-NOW criteria^52^. Furthermore, a class member score of ≥ 0.5 for one of the GBM subclasses was required. Evaluation of the *MGMT* promoter methylation status was made from the classifier output v12.8 using the *MGMT*-STP27 method^53^. All idats corresponding to methylation array data were processed similarly using the minfi package in R (version 1.48.0)^54^. The data were processed using the preprocessIllumina function. Only probes with a detection *P* < 0.01 were kept for further analysis. Also, probes with <3 beads in at least 5% of samples, as well as all non-CpG probes, SNP-related probes, and probes located on X and Y chromosomes were discarded. The CpG intensities were converted to beta values representing total methylation levels (0-the co-registration process was quantified by assessing the correct alignment of ten anatomical landmarks in axial slices (2 x pons, 3^rd^ ventricle, caudate nucleus, lateral ventricle, 2 x central sulcus, superior 1). Missing beta values were imputed with the mean of observed values for CpGs detected in at least 50% of samples. Neurotransmitter receptor methylation was computed as the average beta value across relevant probes. To examine associations between MRI-derived neurotransmitter levels and neurotransmitter receptor DNA methylation, we employed locally weighted scatterplot smoothing (LOWESS) regression. This non-parametric approach was chosen to capture potentially non-linear relationships without assuming a functional form. A smoothing bandwidth of 0.75 was applied, reflecting broad trends across the full range of each neurotransmitter measure. Uncertainty was quantified via a non-parametric bootstrap (1,000 resamples), with pointwise 95% confidence intervals derived from the 2.5th and 97.5th percentiles of the resulting smoothed curves. Analyses were performed in Python (v 3.11.14) using the statsmodels and NumPy libraries.

### Statistical analysis

Statistical comparisons of clinical characteristics between the two datasets were performed using the Student t-test, the Chi-Square Test, or the Fisher test, as appropriate. To compare the magnitude of NT-specific network involvement across the 19 systems, we performed a repeated-measures ANOVA with NT as the within-subject factor using ezANOVA, evaluating sphericity using Mauchly’s test, and applied Greenhouse-Geisser/Huynh-Feldt corrections where appropriate. Post-hoc testing included paired t-tests (171 comparisons), corrected for multiple comparisons using the false discovery rate (FDR), and effect-coded contrasts using the *emmeans* package, comparing each NT score to the grand mean across all NT variables. These contrasts identify which NT systems show the strongest positive or negative deviation from the overall average.

To assess the association between the GBM-related and NT-specific network involvement and OS, we employed a two-step modeling framework that combines (1) Partial Least Squares (PLS) modelling for variable selection and (2) Cox Regression (COXR) proportional hazard modeling for outcome inference and interpretability, adjusted for critical clinical covariates. First, all NT-network scores and Disc-SL were entered into one PLS regression against OS using the *plsr* function in R. Informative predictors were defined based on their cumulative Variable Importance in Projection (VIP) values as a surrogate for each variable’s overall contribution to the model (cutoff VIP>1). Informative predictors were defined based on their cumulative Variable Importance in Projection (VIP) values as a surrogate for each variable’s overall contribution to the model. We retained three latent components in PLS for VIP score computation, following established recommendations that slightly extending the number of components beyond the predictive optimum improves the stability of variable weights and captures distributed covariance patterns in multivariate data^26-29^. To assess the robustness of variable importance, 95% confidence intervals for VIP scores were estimated using 200 repeated subsampling of the dataset. Predictors were considered informative when the lower bound of the 95% confidence interval exceeded 1.0. Only predictors meeting this criterion underwent subsequent outcome inference for interpretation using COXR, adjusted for the following base variables: age, *MGMT* status, and near-GTR. PLS-informed COXR models were compared with base models that confidence intervals are computed for the final models. Analyses were conducted separately in both cohorts, UKE and UPENN. *P*-values of the COXR-derived predictor variables were FDR-corrected for multiple comparisons within each cohort. Winning models, robust across both cohorts, are visualized using Kaplan-Meier curves. For visualization, a median split was performed on NT-specific network involvement, and survival curves are compared between patients with higher and lower NT-network scores. Group differences in OS were assessed using the log-rank test, which compares survival curves by testing the null hypothesis of equal hazard functions over time between groups of higher vs lower NT-network scores. To explore the combined effects of *MGMT* promoter methylation and VAChT network involvement, we subsequently examined differences in overall survival between *MGMT*-methylated and unmethylated tumors and further assessed survival across groups defined by the joint stratification of *MGMT* status and VAChT involvement. This was performed as a joint analysis across the UKE and UPENN cohorts. A two-sided *P*-value<0.05 was considered statistically significant. For sensitivity analyses, the winning model for the UKE cohorts was further explored when adding the administration of radiochemotherapy^23,50^, and the preoperative Karnofsky Performance Score^55^. The best-performing models were subsequently repeated in GBM patients within anatomical regions of interest-the hippocampus, lentiform nucleus, vmPFC, and PFC-to evaluate the anatomical specificity of the identified NT network associations.

### Out-of-sample prediction

To complement the association-based analyses and to explicitly evaluate predictive generalizability, we implemented an out-of-sample prediction framework. For predictive modeling, overall survival was dichotomized using a median split to define the binary outcome. Predictive analyses were performed on the combined UKE and UPENN dataset to maximize statistical power and ensure stable out-of-sample evaluation. Modeling was based on regularized logistic regression with an elastic net penalty, as implemented in the *glmnet package* in R. Elastic net regularization was chosen with joint L1/L2 shrinkage. Predictive models were trained and evaluated using repeated stratified cross-validation with 100 repetitions of 5-fold splitting, ensuring balanced outcome distributions across folds. For each repetition, models were trained on approximately 80% of the data and evaluated on the remaining 20%. To account for outcome imbalance, class-dependent observation weights were applied within each training fold. Hyperparameter tuning for the regularization strength (λ) was performed via inner cross-validation within the training data, and the conservative λ_1_se solution was selected for prediction to promote model stability and parsimony. Two models were evaluated: a base model comprising age, *MGMT* status, and extent of resection, and a full model that additionally incorporated NT-informed network damage measures next to overall network damage (Disc-SL). Predictive performance on held-out test folds was quantified using the area under the receiver operating characteristic curve (ROC-AUC). To characterize predictor contributions within the predictive modeling framework, standardized regression coefficients from the full model at λ_1_se were aggregated across folds and repetitions. This yielded complementary measures of mean absolute coefficient magnitude and selection frequency, thereby assessing both effect strength and robustness under repeated resampling. Together, this approach enabled an included only the base variables using likelihood ratio testing (LRT) and the Akaike Information Criterion (AIC). Hazard ratios (HR) with 95% independent evaluation of the predictive relevance and stability of NT-informed network damage patterns under rigorous covariate adjustment.

## Data Availability

All data produced in the present study are available upon reasonable request to the authors

